# Factors affecting Unplanned Out-of-hospital Births and Neonatal Outcomes at an Urban Hospital

**DOI:** 10.1101/2024.09.09.24313335

**Authors:** Cathy Daichang, Sidney Chu, Helen N Nguyen, Adaora Madubuko

**Affiliations:** Division of Neonatology, Rutgers New Jersey Medical School, Newark, New Jersey, 07103, USA

## Abstract

**Importance:** Unplanned out-of-hospital births have been associated with increased maternal and neonatal complications.

**Objective:** To determine factors that increase unplanned out-of-hospital births incidence and examine neonatal complications.

**Design:** Case-control study, January 2017 to December 2022.

**Setting:** Single-center

**Participants:** Unplanned out-of-hospital births coded under Z38.1 within the hospital database. A random sample of in-hospital births from the same time period served as control. Newborns with chromosomal and congenital abnormalities, stillbirths, and non-singleton births were excluded from analyses.

**Main Outcomes/Measures:** Maternal demographic risk factors for unplanned out-of-hospital births and neonatal complications and morbidity.

**Results:** 66 unplanned out-of-hospital births were compared with 72 in-hospital births. Newborns of unplanned out-of-hospital births were more likely to be affected by low birth weight (OR=5.90, 95%CI [1.87, 18.6]), prematurity (OR=4.84, 95CI% [1.67, 14.1]), and low gestational age compared to in-hospital newborns (p=4.13x10-3). Hypoglycemia (OR=38.0, 95%CI [4.95, 291]), hypothermia (OR=35.5, 95% CI [4.62, 272]), and bradycardia (OR=15.58, 95% CI [0.86, 282]) were significantly associated with unplanned out-of-hospital births. Developmental delay, APGAR scores, and neonatal mortality were not significantly associated with birth location. Black/African-American mothers were significantly more likely to have unplanned out-of-hospital births (OR=4.29, 95%CI [2.10, 8.74]). Mothers with unplanned out-of-hospital births were eight times as likely to have any substance-use-related ICD codes recorded (OR=7.98, 95%CI [2.22, 28.7]) and less likely to receive appropriate prenatal care (OR=0.09, 95%CI [0.03, 0.26]). Maternal parity, age at delivery, marital status, insurance, education, use of interpreting services, and employment status were not significantly associated with birth location.

**Conclusions and Relevance:** Findings suggest that mothers of Black/African-American race or substance-use-related diagnoses are more likely to have unplanned out-of-hospital births and less than appropriate prenatal care. Newborns of this group were more likely to be of low birth weight, low gestational age, and have hypoglycemia, hypothermia, and bradycardia. These findings emphasize the need for targeted interventions for at-risk populations to decrease the risk of preventable neonatal complications.

**KEY POINTS:** *Question:* What are the maternal factors and neonatal outcomes associated with unplanned out-of-hospital births (UOHBs) in an urban, inner-city environment?

*Findings:* This study revealed significant associations to UOHBs that include insufficient prenatal care, substance use disorder, and demographic variables such as race/ethnicity. Newborn outcomes such as preterm births, low birth weight, hypoglycemia, hypothermia and bradycardia were significantly associated with UOHBs.

*Meaning:* These findings emphasize the need for targeted interventions for at-risk populations to decrease the risk of preventable neonatal complications.

## Introduction

Unplanned out-of-hospital births (UOHB), when an infant is unintentionally born outside a hospital setting^1^, should be differentiated from planned out-of-hospital births that were intended to occur at home or at a freestanding birth center.^2^ UOHB have previously been associated with including neonatal hypothermia, hypoglycemia, polycythemia^3^, increased neonatal mortality^3^, and admission to the neonatal intensive care unit due to complications.^4^ Another study found that maternal diabetes mellitus, hypertensive disorders of pregnancy, and obesity were associated with poor perinatal mortality outcomes in the setting of UOHB.^5^ However, few studies have examined long-term health outcomes in children of UOHB, although, there have been studies on neonatal mortality rates in planned at-home births versus in-hospital births.^6^

Previous studies showed history of cocaine use, lack of health insurance^3^, inadequate prenatal care^5^, maternal age over 35, high parity, and lower educational attainment^4^ as potential risk factors for UOHB. Moreover, one study found that women from lower socioeconomic backgrounds had increased odds of having an UOHB.^7^ This study seeks to understand risk factors and outcomes of UOHB at University Hospital in Newark, NJ, which has a poverty rate more than twice the national average (24.44% vs 11.5%, respectively).^8^ Non-Hispanic black race and poverty showed significant impact on infant mortality in Newark.^9^ From 2017-2022, the rate of out-of-hospital deliveries at University Hospital was 10.4 births per 1000 births over the period of study (1.04%). and about four times greater than the national rate of about 0.25% (eFigure 1).^10,11^ Examining maternal risk factors and newborn outcomes of unplanned out-of-hospital births at University Hospital will provide insight into improving clinical outcomes for the most at-risk infants and mothers.

**Figure 1.**
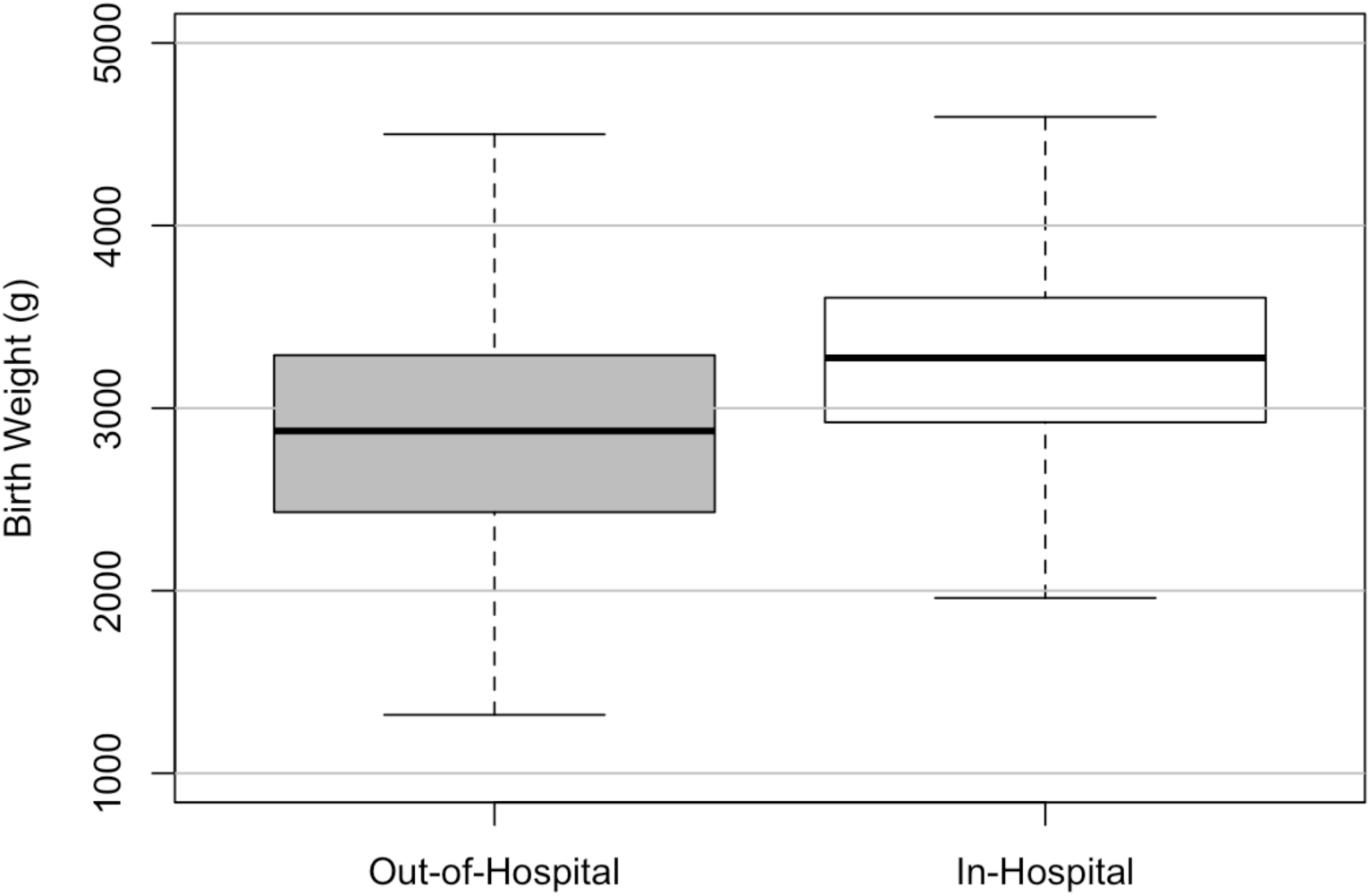
Comparison of Birth Weight by Birth Location. Gray box represents out-of-hospital birth group, white box represents in-hospital birth group. Birth weight in grams (g) is represented on the vertical axis. Both boxes represents the interquartile range (IQR) of birth weights of neonates in their respective birth location. Median birth weight is shown by the horizontal line within the box. Whiskers extend to the minimum and maximum birth weights within 1.5 times the IQR.

## Methods

### Study Design

This observational study was conducted at University Hospital (Newark, New Jersey, USA) with IRB approval; patient data from January 2017 to December 2022 was retroactively sampled through Epic electronic medical record (EMR) Both UOHB and in-hospital birth groups were stratified into maternal and neonatal cases and studied independently.

### Sample Size and Subject Selection

66 unplanned out-of-hospital live singleton births and 72 in-hospital live singleton births within the study period were used for analysis. The UOHB group was identified through an EMR-wide search using the code Z38.1, while the control group of in-hospital live singleton births was randomly selected, including neonates with the last digit of 5 in their medical record number, along with their corresponding mothers. Cases with chromosomal abnormalities, stillbirths, non-singleton births, and congenital neonatal anomalies were excluded.

### Data Collection

Via retrospective search of medical records from each individual patient, maternal and neonatal categorical inputs were derived from patient chart files (discharge summaries, history and physical notes, labor and delivery notes, procedure notes, internal communications) while removing patient-identifying information. Evaluation of medical records for maternal characteristics (Table 1) included: 1) age at delivery, 2) parity, 3) marital status, 4) race/ethnicity, 5) primary language, 6) utilization of interpreting services, 7) whether a primary care provider was listed in chart, 8) insurance coverage, 9) substance use history, 10) complications related to pregnancy or any associated diagnosis codes within the prenatal period, 11) educational attainment, 12) employment status, and 13) number of prenatal visits.

**Table 1.**
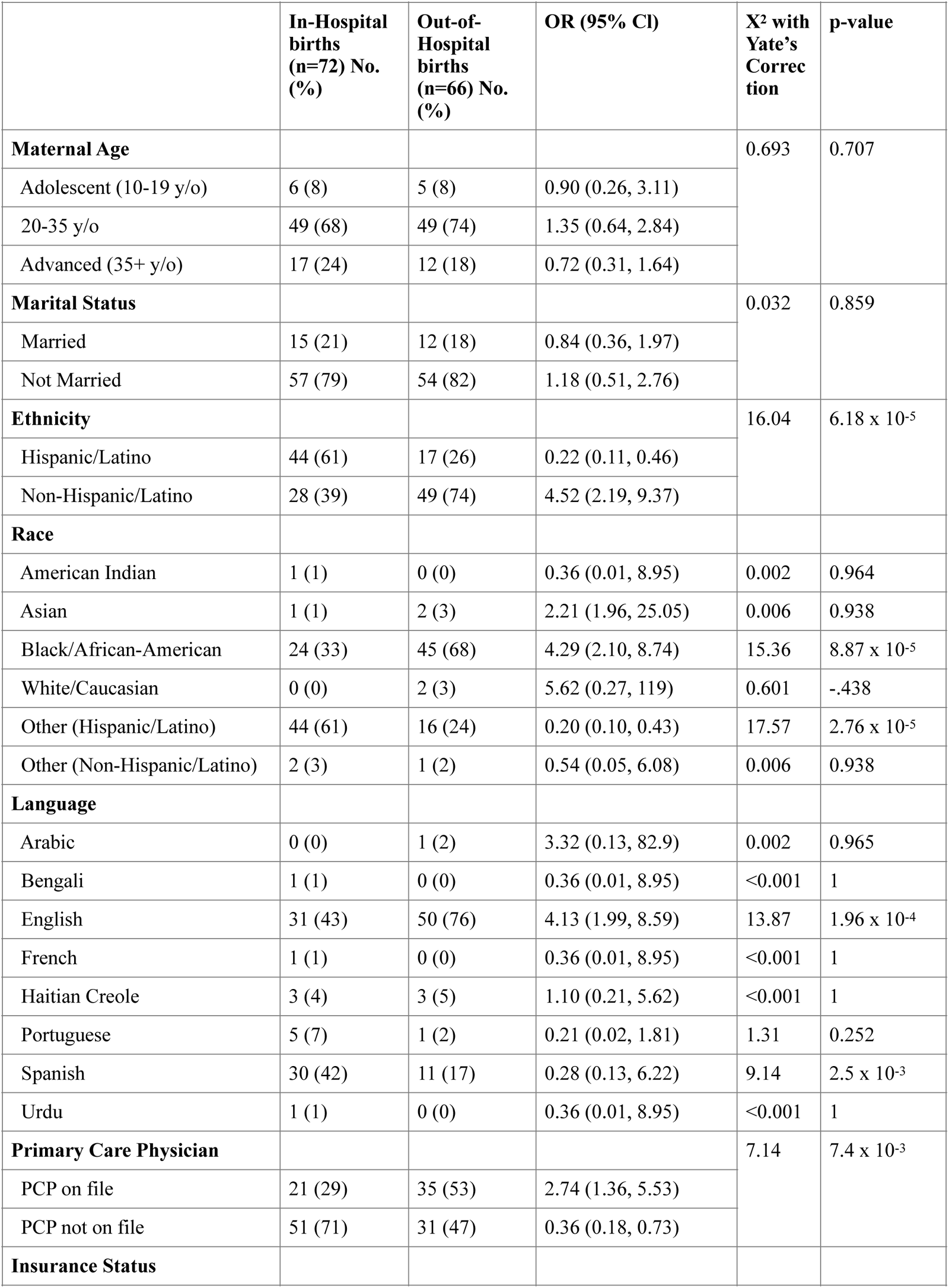
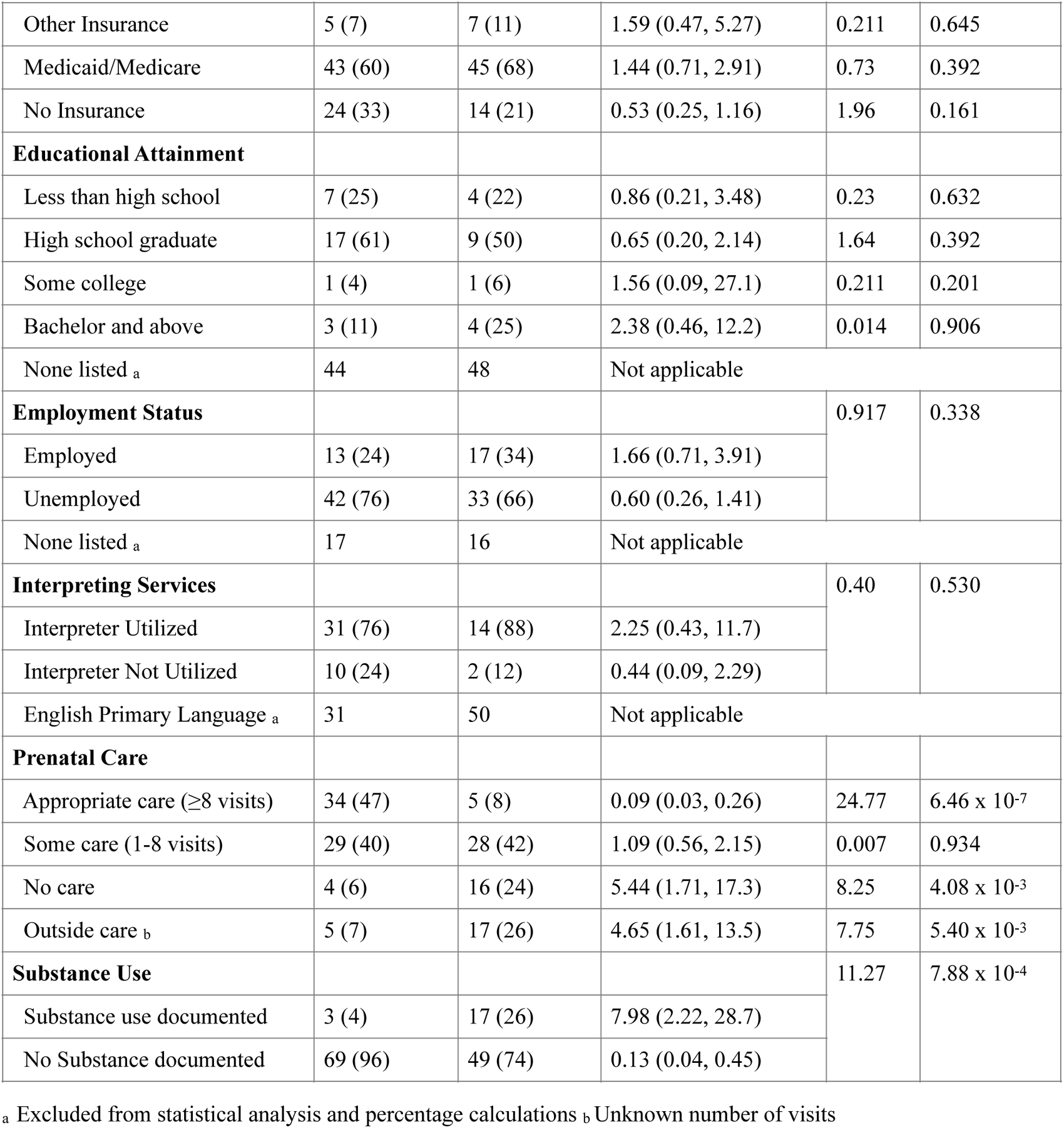
Maternal Categorical Sociodemographic Characteristics.

|  | **In-Hospital**<br>**births**<br>**(n=72) No.**<br>**(%)** | **Out-of-**<br>**Hospital**<br>**births**<br>**(n=66) No.**<br>**(%)** | **OR (95% CI)** | **X<sup>2</sup> with**<br>**Yate's**<br>**Correc**<br>**tion** | **p-value** |
| --- | --- | --- | --- | --- | --- |
| **Maternal Age** |  |  |  | 0.693 | 0.707 |
| Adolescent (10-19 y/o) | 6 (8) | 5 (8) | 0.90 (0.26, 3.11) |  |  |
| 20-35 y/o | 49 (68) | 49 (74) | 1.35 (0.64, 2.84) |  |  |
| Advanced (35+ y/o) | 17 (24) | 12 (18) | 0.72 (0.31, 1.64) |  |  |
| **Marital Status** |  |  |  | 0.032 | 0.859 |
| Married | 15 (21) | 12 (18) | 0.84 (0.36, 1.97) |  |  |
| Not Married | 57 (79) | 54 (82) | 1.18 (0.51, 2.76) |  |  |
| **Ethnicity** |  |  |  | 16.04 | 6.18 x 10<sup>-5</sup> |
| Hispanic/Latino | 44 (61) | 17 (26) | 0.22 (0.11, 0.46) |  |  |
| Non-Hispanic/Latino | 28 (39) | 49 (74) | 4.52 (2.19, 9.37) |  |  |
| **Race** |  |  |  |  |  |
| American Indian | 1 (1) | 0 (0) | 0.36 (0.01, 8.95) | 0.002 | 0.964 |
| Asian | 1 (1) | 2 (3) | 2.21 (1.96, 25.05) | 0.006 | 0.938 |
| Black/African-American | 24 (33) | 45 (68) | 4.29 (2.10, 8.74) | 15.36 | 8.87 x 10<sup>-5</sup> |
| White/Caucasian | 0 (0) | 2 (3) | 5.62 (0.27, 119) | 0.601 | -.438 |
| Other (Hispanic/Latino) | 44 (61) | 16 (24) | 0.20 (0.10, 0.43) | 17.57 | 2.76 x 10<sup>-5</sup> |
| Other (Non-Hispanic/Latino) | 2 (3) | 1 (2) | 0.54 (0.05, 6.08) | 0.006 | 0.938 |
| **Language** |  |  |  |  |  |
| Arabic | 0 (0) | 1 (2) | 3.32 (0.13, 82.9) | 0.002 | 0.965 |
| Bengali | 1 (1) | 0 (0) | 0.36 (0.01, 8.95) | <0.001 | 1 |
| English | 31 (43) | 50 (76) | 4.13 (1.99, 8.59) | 13.87 | 1.96 x 10<sup>-4</sup> |
| French | 1 (1) | 0 (0) | 0.36 (0.01, 8.95) | <0.001 | 1 |
| Haitian Creole | 3 (4) | 3 (5) | 1.10 (0.21, 5.62) | <0.001 | 1 |
| Portuguese | 5 (7) | 1 (2) | 0.21 (0.02, 1.81) | 1.31 | 0.252 |
| Spanish | 30 (42) | 11 (17) | 0.28 (0.13, 6.22) | 9.14 | 2.5 x 10<sup>-3</sup> |
| Urdu | 1 (1) | 0 (0) | 0.36 (0.01, 8.95) | <0.001 | 1 |
| **Primary Care Physician** |  |  |  | 7.14 | 7.4 x 10<sup>-3</sup> |
| PCP on file | 21 (29) | 35 (53) | 2.74 (1.36, 5.53) |  |  |
| PCP not on file | 51 (71) | 31 (47) | 0.36 (0.18, 0.73) |  |  |
| **Insurance Status** |  |  |  |  |  |
| Other Insurance | 5 (7) | 7 (11) | 1.59 (0.47, 5.27) | 0.211 | 0.645 |
| Medicaid/Medicare | 43 (60) | 45 (68) | 1.44 (0.71, 2.91) | 0.73 | 0.392 |
| No Insurance | 24 (33) | 14 (21) | 0.53 (0.25, 1.16) | 1.96 | 0.161 |
| **Educational Attainment** |  |  |  |  |  |
| Less than high school | 7 (25) | 4 (22) | 0.86 (0.21, 3.48) | 0.23 | 0.632 |
| High school graduate | 17 (61) | 9 (50) | 0.65 (0.20, 2.14) | 1.64 | 0.392 |
| Some college | 1 (4) | 1 (6) | 1.56 (0.09, 27.1) | 0.211 | 0.201 |
| Bachelor and above | 3 (11) | 4 (25) | 2.38 (0.46, 12.2) | 0.014 | 0.906 |
| None listed <sub>a</sub> | 44 | 48 | Not applicable |  |  |
| **Employment Status** |  |  |  | 0.917 | 0.338 |
| Employed | 13 (24) | 17 (34) | 1.66 (0.71, 3.91) |  |  |
| Unemployed | 42 (76) | 33 (66) | 0.60 (0.26, 1.41) |  |  |
| None listed <sub>a</sub> | 17 | 16 | Not applicable |  |  |
| **Interpreting Services** |  |  |  | 0.40 | 0.530 |
| Interpreter Utilized | 31 (76) | 14 (88) | 2.25 (0.43, 11.7) |  |  |
| Interpreter Not Utilized | 10 (24) | 2 (12) | 0.44 (0.09, 2.29) |  |  |
| English Primary Language <sub>a</sub> | 31 | 50 | Not applicable |  |  |
| **Prenatal Care** |  |  |  |  |  |
| Appropriate care (≥8 visits) | 34 (47) | 5 (8) | 0.09 (0.03, 0.26) | 24.77 | 6.46 x 10<sup>-7</sup> |
| Some care (1-8 visits) | 29 (40) | 28 (42) | 1.09 (0.56, 2.15) | 0.007 | 0.934 |
| No care | 4 (6) | 16 (24) | 5.44 (1.71, 17.3) | 8.25 | 4.08 x 10<sup>-3</sup> |
| Outside care <sub>b</sub> | 5 (7) | 17 (26) | 4.65 (1.61, 13.5) | 7.75 | 5.40 x 10<sup>-3</sup> |
| **Substance Use** |  |  |  | 11.27 | 7.88 x 10<sup>-4</sup> |
| Substance use documented | 3 (4) | 17 (26) | 7.98 (2.22, 28.7) |  |  |
| No Substance documented | 69 (96) | 49 (74) | 0.13 (0.04, 0.45) |  |  |

<sup>a</sup> Excluded from statistical analysis and percentage calculations <sup>b</sup> Unknown number of visits

Evaluation of medical records for neonatal characteristics (Table 2) included: 1) birth weight^12^, 2) gestational age at birth (in weeks and days) and preterm status (< 37 weeks as defined by the World Health Organization^13^), 3) 5-minute APGAR score, 4) chart record of neonatal survival (0-28 days since birth) and perinatal survival (29-154 days since birth), 6) complications (Table 3), and 7) record of developmental delay at 18-month check-up.

**Table 2.** Neonatal Categorical Characteristics.

|  | **In-Hospital**<br>**births**<br>**(n=72) No.**<br>**(%)** | **Out-of-**<br>**Hospital**<br>**births**<br>**(n=66) No.**<br>**(%)** | **OR (95% CI)** | **X<sup>2</sup> or**<br>**Fisher's**<br>**Exact Test** | **p-value** |
| --- | --- | --- | --- | --- | --- |
| **Birth Weight** |  |  |  |  |  |
| < 2500 grams | 4 (6) | 17 (26) | 5.90 (1.87, 18.6) | 9.38<sub>a</sub> | 2.19 x 10<sup>-3</sup> |
| ≥ 2500 – 4000 grams | 64 (88) | 47 (71) |  | 5.76<sub>a</sub> | 1.63 x 10<sup>-2</sup> |
| ≥4000g | 4 (6) | 2 (3) |  | 0.095<sub>a</sub> | 0.757 |
| **Gestational Age at Birth** |  |  |  | 8.22<sub>a</sub> | 4.13 x 10<sup>-3</sup> |
| Full term (> 37w) | 67 (93) | 47 (73) | 0.20 (0.07, 0.60) |  |  |
| Pre-term (< 37w) | 5 (7) | 17 (27) | 4.84 (1.67, 14.1) |  |  |
| Unknown gestational age <sub>c</sub> | 0 | 2 |  |  |  |
| **Developmental Delay at 18 months** |  |  |  | 0.120<sub>a</sub> | 0.729 |
| Present | 4 (14) | 2 (18) | 1.39 (0.21, 8.93) |  |  |
| Absent | 25 (86) | 9 (82) | 0.72 (0.11, 4.63) |  |  |
| No follow up recorded <sub>c</sub> | 43 | 55 |  |  |  |
| **Neonatal Survival** |  |  |  | Not applicable<sub>b</sub> | 0.4095 |
| Survival | 62 (100) | 42 (98) | 0.23 (0.01, 5.70) |  |  |
| Non-Survival | 0 (0) | 1 (2) | 4.41 (0.18, 111.0) |  |  |
| No follow up recorded <sub>c</sub> | 10 | 23 |  |  |  |

<sub>a</sub> Calculated with X<sup>2</sup> with Yate's Correction <sub>b</sub> Calculated with Fisher's Exact Test <sub>c</sub> Excluded from statistical analysis and percentage calculations

**Table 3.** Neonatal Complications.

|  | **In-Hospital**<br>**births**<br>**(n=72) No.**<br>**(%)** | **Out-of-**<br>**Hospital**<br>**births**<br>**(n=66) No.**<br>**(%)** | **OR (95% CI)** | **X<sup>2</sup> with**<br>**Yate's**<br>**Correction** | **p-value** |
| --- | --- | --- | --- | --- | --- |
| Polycythemia | 0 (0) | 5 (8) | 12.97 (0.70, 239) | 3.70 | 0.054 |
| Hypoglycemia | 1 (1) | 23 (35) | 38.0 (4.95, 291) | 24.56 | 7.22 x 10<sup>-7</sup> |
| Convulsions | 1 (1) | 0 (0) | 0.36 (0.01, 8.95) | < .001 | 1 |
| Hypothermia | 1 (1) | 22 (33) | 35.5 (4.62, 272) | 23.05 | 1.58 x 10<sup>-6</sup> |
| Bradycardia | 0 (0) | 6 (9) | 15.58 (0.86, 282) | 4.83 | 2.79 x 10<sup>-2</sup> |
| Respiratory Distress | 10 (14) | 14 (21) | 1.67 (0.68, 4.07) | 0.83 | 0.36 |
| Persistent Pulmonary<br>Hypertension of the Newborn | 1 (1) | 5 (8) | 5.81 (0.66, 51.2) | 1.86 | 0.17 |
| Metabolic or Respiratory<br>Acidosis | 3 (4) | 3 (5) | 1.10 (0.21, 5.63) | < .001 | 1 |

### Statistical Analysis

Standard statistical tests (Welch’s two sample t-test, chi-square test of independence with Yates’ correction, and Fisher’s exact test) were applied in comparing in-hospital birth groups with the UOHB group. Two-tailed p-values ≤0.05 were considered statistically significant. In both maternal and newborn analyses, chi-square tests of independence based on 2x2 contingency tables stratified by each dichotomous independent variable and birth group/location.Yates’ correction was used in chi-square tests to reduce type I error. For analyses with multiple variable subsets like race (6) or birth weight (3), separate 2x2 contingency tables were used for each sub-category. Fisher’s exact test was applied in mortality/survival analysis to handle low counts conservatively. Number of prenatal care visits and birth weight analysis utilized Welch’s two-sample t-test and a boxplot for illustration.

## Results

The UOHB trend at University Hospital appeared stable throughout the study period, with a notable increase in 2022. The COVID pandemic (2019-2021) did not significantly affect out-of-hospital birth rates (eFigure 2).

**Figure 2.**
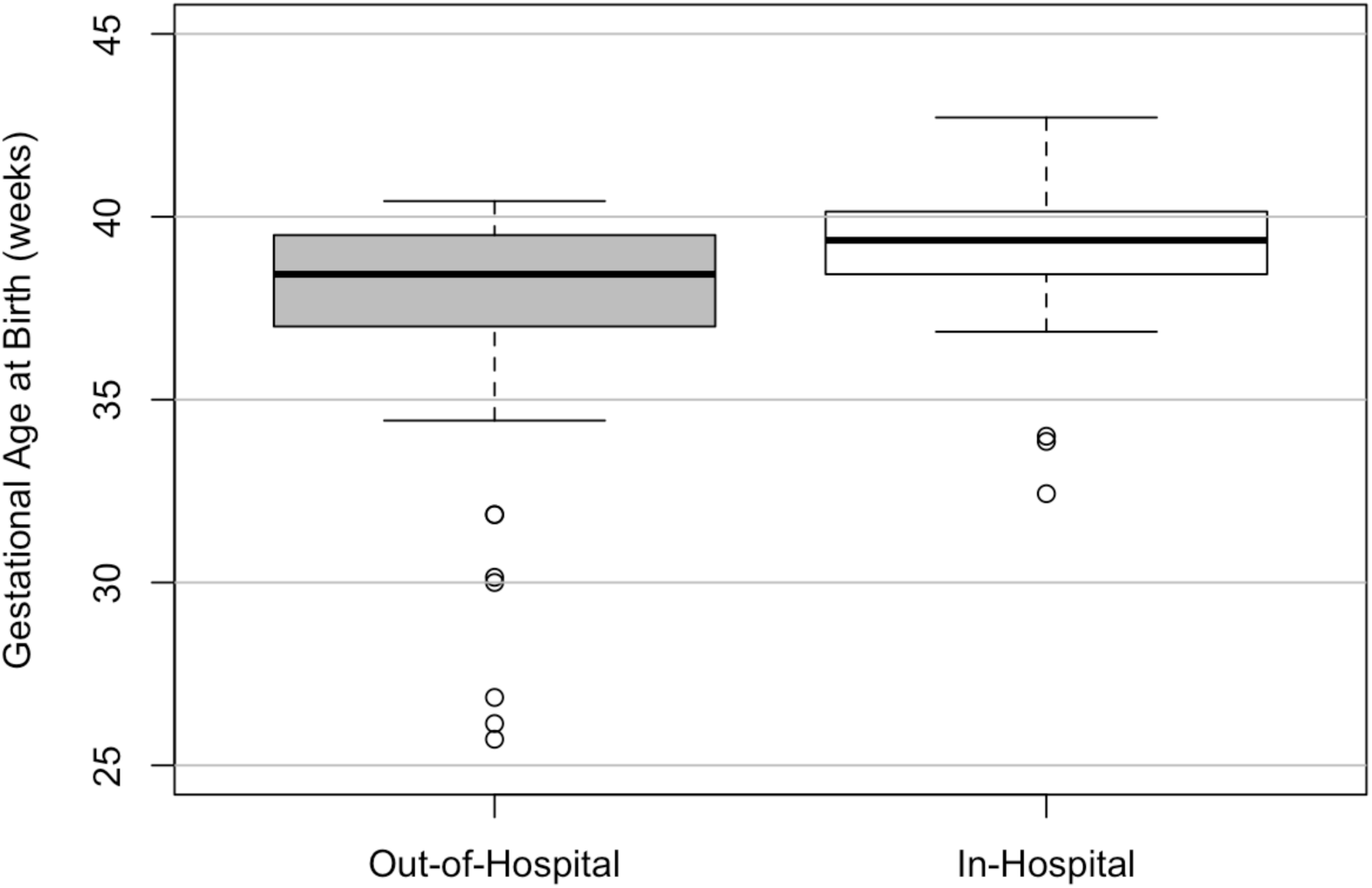
Comparison of Gestational Age by Birth Location. Gray box represents out-of-hospital birth group, white box represents in-hospital birth group. Gestational age in weeks is represented on the vertical axis. Both boxes represents the interquartile range (IQR) of the gestational age at birth of neonates in their respective birth location. Median gestational age at birth is shown by the horizontal line within the box. Whiskers extend to the minimum and maximum birth weights within 1.5 times the IQR. Circles represent neonates whose gestational ages fall beyond 1.5 times the IQR.

### Maternal Demographics and Characteristics

Out-of-hospital and in-hospital groups revealed similar distributions in maternal age at delivery; the mean age at delivery in the out-of-hospital group was 27.9 and the mean age at delivery in the in-hospital group was 27.7. There was no significant difference between the true averages of maternal age at delivery between both groups (p=0.875) or maternal age categories and birth location (p=0.707) (Table 1). The mean parity in the out-of-hospital group was 2.29, while the mean parity in the in-hospital group was 2.01. A Welch’s two sample T-test revealed no significant difference of true mean in both locations (p=.875, [-0.84, 0.287]).

Patients who identified as Black/African-American were significantly more likely to have UOHB (p=8.87 x 10^-5^, OR=4.29 [2.10, 8.74]), while Hispanic/Latino patients were significantly less likely to have UOHB (p=2.76x10^-5^, OR=0.20 [0.10, 0.43]) All other categories were not significantly associated with birth location; the Pacific Islander and Mixed categories were excluded due to lack of data. In a separate analysis stratified by ethnicity alone, Hispanic/Latino ethnicity was significantly negatively associated with UOHB (p=6.18 x 10^-5^, OR=0.22 [0.11, 0.46]) (Table 1).

Mothers whose primary language was English were significantly more likely to have UOHB (p=1.96 x 10^-4^, OR=4.13 [1.99, 8.59]), whereas mothers whose primary language was Spanish were significantly less likely to have UOHB (p=2.50x10^-3^, OR=0.28 [0.13, 6.22]). All other languages demonstrated no significant association with birth location (Table 1). When analyzing patients whose primary language was not English (n=57), there was no significant association between interpreting services utilization and birth location (p=0.530) (Table 1).

Having a primary care provider on-file significantly increased the odds of having an UOHB (OR=2.74 [1.36, 5.53]). Independently, there was no significant association between educational attainment, employment status, insurance coverage, or marital status and birth location (Table 1).

UOHB were significantly less likely to receive adequate prenatal care (OR=0.09 [0.03, 0.26]) and were more likely to receive outside (OR=4.65 [1.61, 13.5]) or no prenatal care at all (OR=5.44 [1.71, 17.3]) (Table 1). Of the UOHB group, only 8% received adequate prenatal care (n=5). A significant difference (p=3.51x10 -9, 95%CI [-5.33, -2.81]) was observed in prenatal visits between the two groups. Excluding outside prenatal care, in-hospital births had an average of 7.36 prenatal visits, while UOHB had an average of 3.28 visits.

Substance use was found to be significantly associated with birth location (p=7.88 x 10^-4^), with mothers with documented substance use during pregnancy about 8 times as likely to give birth out of hospital (OR=7.98 [2.22, 28.7]) (Table 1).

### Neonatal Characteristics and Complications

Out-of-hospital births were almost six times as likely to be low birth weight (OR=5.90, 95%CI [1.87, 18.6]) (Table 2). In-hospital births were more likely to achieve normal birth weight (*p*=1.63x10^-2^), and less likely to be low birth weight (*p*=2.19 x 10^-3^). Macrosomia was not significantly associated with birth location. There was a significant difference (*p*=8.76 x 10^-5^) between the birth weight of both groups; the mean birth weight in the in-hospital group was 3269g while the mean birth weight in the UOHB group was 2850g (Figure 1).

There was a significant association (*p*=4.13x10^-3^) between gestational term and birth location; out-of-hospital births were almost five times as likely (OR=4.84, 95%CI [1.67, 14.1]) to be born preterm (Table 2). Excluding those with unknown gestational age, there was a significant difference in gestational age of birth between both groups (*p*=4.45 x 10^-4^): the mean gestational age at birth in the in-hospital group was 39 2/7 weeks versus the UOHB group at 38 3/7 weeks (Figure 2).

Many UOHB did not have recorded APGAR scores and only 28 cases were included for this analysis. Given this, there was no significant difference (*p*=0.8912) between the out-of-hospital and in-hospital group. Hypoglycemia (*p*=7.22x10^-7^), hypothermia (*p*=1.58x10^-6^) and bradycardia (*p*=2.79x10^-2^) were significantly associated with UOHB. Out-of-hospital neonates were 38 times more likely to suffer from hypoglycemia (OR=38.0, 95%CI [4.95, 291]), 35 times more likely to suffer from hypothermia (OR=35.5, 95%CI [4.62, 272]) and 15 times more likely to suffer from bradycardia (OR=15.58, 95%CI [0.86, 282]). Polycythemia, convulsions, respiratory distress, PPHN, and metabolic/respiratory acidosis were not significantly associated with birth location (Table 3).

### Infant Characteristics: Long-Term Consequences

As only one non-survival count was recorded in the neonatal period, Fisher’s exact test was employed: there was no significant association between neonatal survival and birth location (Table 2). Perinatal survival analysis was excluded due to lack of non-survival counts.

Developmental delay was defined as failure to reach any number of developmental milestones by 18 months of age or any other associated developmental pathology. A majority of patients were lost to follow-up before their 18-month check-up and were excluded from analysis. No significant association was found between developmental delay and birth location (Table 2).

## Discussion

### Maternal Characteristics

In this study, no significant correlation was found between maternal age and birth location in either the adolescent or advanced age groups, while the mean maternal ages at delivery for both the out-of-hospital and in-hospital groups (27.9 and 27.7, respectively) reveals a striking proximity to the national average age of mothers at first birth, reported as 27.3 years by the CDC for the year 2020.^14^ This differs from previous studies emphasizing maternal age over 35 as a risk factor for UOHB.^4^ One possible reason is that our sample had a higher percentage of patients under 20 years of age (7.2%) compared to a national average of 4.4% from 2019-2021^15^ and NJ average of 2.4% in 2021.^16^ Additionally, only 16% in our sample were 35 or older, compared to 19% nationally.^17^ This suggests our sample skewed younger; with a small sample, definitive conclusions on the maternal age-UOHB link are challenging. Further investigation with larger samples is recommended.

Parity was not significantly associated with unplanned out-of-hospital deliveries, while other studies found that multiparous women were at risk for UOHB.^5,18^ This discrepancy may be influenced by factors discussed earlier (age and birth location) and warrants further research for clarification.

The observed racial disparities in birth location, especially the increased likelihood of UOHB among Black/African-American mothers, are consistent with existing literature.^19^ This disparity is linked to higher maternal mortality rates and adverse birth outcomes among Black women, influenced by systemic racism, socio-economic disparities, and limited healthcare access.^20,21^ Notably, in-hospital maternal mortality is over twice as high among Black patients compared to White patients.^22^ New Jersey’s maternal mortality rate, although high, has seen some improvement, particularly benefiting Black/African American mothers who are nearly seven times more likely than white mothers in the state to die from maternity-related complications.^23^ Research has found that poverty is associated with a preference for out-of-hospital births, emphasizing these disparities.^24^

In contrast, “other race” (Hispanic/Latino) mothers were significantly less likely to have UOHB. When stratified by ethnicity alone, Hispanic/Latino mothers were, again, significantly less likely to have UOHB. These findings align with a 2022 report from the CDC showing that Hispanic mothers have the lowest percentage of home births compared to White or Black Non-Hispanic mothers, although the percentage has increased each year since 2019.^25^ Further study is recommended to examine both the protective factors against UOHB and the factors leading to an increasing UOHB rate in this group.

The association between language and birth location is a unique aspect of this study; while there is limited research directly exploring this relationship, previous research associated language barriers with reduced access to healthcare services, poorer health outcomes, and increased healthcare disparities.^26^ The higher likelihood of UOHB among English-speaking mothers compared to non-English-speaking mothers seems to contradict these known associations. This may be explained by the unique demographic characteristics of the study population: a significant portion of Hispanic/Latino patients were recent immigrants who were non-English-speaking rather than those of Hispanic/Latino origin who spoke English. The lower likelihood of UOHB among Hispanic/Latino mothers in this study could contribute to the observed language disparity.

Mothers with UOHB were almost eight times as likely to have any substance-use-related ICD code recorded in maternal charts. Substance use during pregnancy is consistently linked to adverse birth outcomes, including preterm birth and low birth weight.^27^ Given this, there is a need to distinguish between mothers with documented substance use during pregnancy and those who were actively under the influence during their UOHB, although incomplete documentation limited further stratification. Some cases involved mothers unaware of labor due to substance effects and unable to transport themselves to a hospital. These results underscore the need for comprehensive maternal care addressing substance use disorders, including counseling pregnant women on contingency plans and safeguards during labor. Reporting bias may confound these findings, as healthcare providers may be more likely to inquire about substance use in cases of UOHB.

Having a primary care physician (PCP) on-file was significantly associated with having an UOHB. This may be due to lack of consistency in obtaining this information from patients, though more insight is needed. One possibility is that when presented with an UOHB with little prenatal care, providers will more regularly inquire about and document PCPs in the EMR to obtain medical information that may otherwise be difficult to access. Additionally, while educational attainment, employment status, insurance coverage, or marital status were not correlated with birth location, data on these factors were frequently not listed in patient charts. This emphasizes the need for thorough medical documentation to be able to retroactively study this patient group.

The stark difference of the average of 3.3 prenatal visits in UOHB versus 7.3 visits in the control group suggests that the few prenatal visits UOHB mothers tend to have could be a powerful intervention point to educate patients on the risks of UOHB and warning signs of labor. If a provider notices that a patient with the risk factors identified above is lacking or falling behind on their prenatal visits, preemptive counseling could prevent the negative health outcomes of UOHB.

### Neonatal Characteristics and Complications

Preterm births were significantly associated with out-of-hospital births, which aligns with existing literature.^5^ The disparity in preterm birth rates for Black women in the US may underscore this association.^28^ Moreover, births with unknown gestational ages were excluded from analysis, which could indicate that the association could theoretically be even stronger as UOHB had significantly less prenatal care and could be more likely to have unknown gestational ages. This increased preterm birth rate of UOHB is likely to explain the other significant association of lower birth weight and UOHB.

The observed association between UOHB and adverse neonatal outcomes, including hypoglycemia, hypothermia, and bradycardia, underscores the vulnerability of infants born unintentionally outside a healthcare facility. These findings align with existing literature, emphasizing the heightened risks for UOHB newborns, especially in urban settings. Out-of-hospital newborns may face increased hypothermia risk due to limited fat and glycogen reserves for temperature control in cold environments or if not dried properly, particularly if premature or with low birth weight.^29,30^ The increased occurrence of these complications highlights the importance of monitoring all out-of-hospital births for hypoglycemia, hypothermia, and bradycardia.

While these complications can often be prevented through standard in-hospital care practices, such as infant warming, drying, and controlled umbilical cord clamping^30,31^, a limitation of this analysis is the challenge in distinguishing whether the increased rates of these complications are due to prematurity, being born out-of-hospital, or a combination of both. Without a study comparing neonates with similar gestational age and birth weight from both groups, it is difficult to attribute these complications solely to being an UOHB. Nevertheless, from a clinical perspective, this distinction has limited value; the strong associations between these complications and UOHB should guide providers in the initial treatment of UOHB neonates.

There was no significant association between developmental delay or neonatal mortality and birth location in the follow-up analysis; statistical power was limited by high rates of loss to follow-up. Future research with larger samples could provide insights into the long-term developmental outcomes of infants of UOHB.

### General Limitations

Because this study is hospital-based rather than population-based, conclusions most directly apply to patients in a single inner-city hospital and may not be generalizable to other areas. The hospital’s distinct patient population introduces confounding factors and may lead to missed correlations due to inadequate patient data and documentation. To establish broader, conclusive insights, further research should encompass diverse patient populations.

## Conclusion

This study examined multiple interconnected associations to UOHB that include insufficient prenatal care, substance use disorder, and demographic variables such as race/ ethnicity. UOHB were significantly associated with preterm births, low birth weight, hypoglycemia, hypothermia and bradycardia. These findings emphasize the need for targeted interventions for at-risk populations to decrease the risk of preventable neonatal complications from UOHB.

## Data Availability

All data produced in the present study are available upon reasonable request to the authors.

